# *Plasmodium falciparum* artemisinin-resistant K13 mutations confer a sexual-stage transmission advantage that can be overcome with atovaquone-proguanil

**DOI:** 10.1101/2020.10.26.20214619

**Authors:** Zuleima Pava, Sachel Mok, Katharine A. Collins, Maria Rebelo, Rebecca E. Watts, Gregory J. Robinson, Claire Y.T. Wang, Hayley Mitchell, Sean Lynch, Jeremy Gower, Lachlan Webb, Sam McEwan, Anand Odedra, Bridget Barber, Louise Marquart, Matthew W.A. Dixon, Joerg J. Moehrle, David A. Fidock, James S. McCarthy

**Author notes:** **MATERIALS & CORRESPONDENCE** Correspondence to Zuleima Pava at; James McCarthy at; David Fidock at and Sachel Mok at.

## Abstract

Containing the spread of artemisinin (ART)-resistant *Plasmodium falciparum* will be assisted by improved understanding of its human-to-mosquito transmission. We compared gametocyte dynamics among field isolates containing K13 mutations conferring ART resistance and K13 wild-type parasites. In Pailin, Cambodia, the male to female gametocyte ratio was higher among K13 mutant infections compared to K13 wild-type infections. We also investigated the effects of artesunate and atovaquone-proguanil on the transmissibility of an ART-resistant K13 mutant strain, Cam3.II^R539T^, in a volunteer infection study. Gametocyte production was higher after a single dose of artesunate (2 mg/kg) in volunteers infected with ART-resistant compared to ART-sensitive parasites. Despite the presence of gametocytes in volunteers infected with ART-resistant parasites, there was no infection observed in *Anopheles stephensi* mosquitoes after atovaquone-proguanil treatment. We report transmission determinants of ART-resistant infections that could be advantageous over ART-sensitive infections. Moreover, we show additional benefits of treating ART-resistant infections with atovaquone-proguanil treatment.

## INTRODUCTION

The appearance and spread of *Plasmodium falciparum* strains resistant to artemisinin (ART) derivatives, the main component of first-line antimalarial therapies, threatens malaria control and elimination. Infections caused by ART-resistant parasites are characterised by slower parasite clearance after treatment and the presence of point mutations such as C580Y, Y493H, and R539T in the beta-propeller domain of the *P. falciparum* Kelch13 (K13) protein (*PF3D7_1343700*)^1^. ART-resistant infections were first reported in western Cambodia and the Thailand–Cambodia border in 2002–2004^2^. Over the past 10 to 15 years, ART-resistant parasites have come to dominate across the Greater Mekong Subregion^1^. Recently, K13 mutations associated with ART resistance have also been detected in other areas such as Guyana, Papua New Guinea and Rwanda^3-5^.

Higher levels of gametocytes - the parasite stage infectious to mosquitoes - have been suggested among patients infected with ART-resistant *P. falciparum* parasites^1^. These resistant parasites might, therefore, have higher transmissibility. However, because gametocyte levels can be affected by different variables including genetic background^6^, accumulation of gametocytes after a long asymptomatic infection^7^, and antimalarial drug exposure^8-11^, a thorough examination of gametocyte dynamics in ART-resistant infections is needed.

In addition, antimalarials with transmission-blocking activities against ART-resistant strains are needed to slow down or stop their spread. Detailed investigation of the effects of antimalarials on gametocyte dynamics, using data from field studies, has been hampered by several limitations. These include the inability to ascertain the duration of infections prior to individuals seeking treatment, the difficulty of directly estimating the sexual commitment rate (the number of young gametocytes produced per asexual parasite cycle), and the paucity of data on male to female gametocyte ratios^12^.

Volunteer infection studies are playing an increasingly important role in accelerating antimalarial drug development, and in improving our understanding of *P. falciparum* gametocyte dynamics^13-15^. In the Induced Blood Stage Malaria (IBSM) model, healthy volunteers are inoculated intravenously with blood-stage *Plasmodium* parasites and monitored closely using molecular assays. In addition, xenodiagnosis by direct skin feeding or membrane feeding assays, using blood samples collected from infected volunteers, can be performed to determine infectiousness to mosquitoes^13,16^. Recently, we have reported an IBSM model using the ART-resistant *P. falciparum* K13 mutant Cam3.II^R539T^ strain^17^.

Herein, we used data from two complementary studies to test for differences in gametocyte dynamics, including male to female gametocyte ratios and gametocyte levels, in patients and volunteers infected with K13 mutant and K13 wild-type *P. falciparum* parasites. We have also leveraged the ART-resistant IBSM model to compare the effect of a single dose of artesunate on gametocyte production in volunteers infected with ART-resistant or ART-sensitive strains and to investigate the effect of atovaquone-proguanil (AP) treatment on the transmissibility of ART-resistant parasites.

## RESULTS

### Variation in gametocyte dynamics across field sites from the TRAC study

We used the mean log_2_ expression ratio of 121 late-stage gametocyte-specific genes to determine gametocyte levels in 771 clinical samples collected from 11 field sites, as this strongly correlated with the ratio of gametocyte to total parasite numbers as counted by microscopy across all sites (r=0.73, *p*<0.0001) (Supplementary Fig 1A). Gametocyte transcript levels varied significantly among field sites (Kruskal-Wallis test, *p*<0.0001) (Fig. 1A). Samples collected from Pailin, Cambodia had statistically significant higher levels of gametocyte transcripts than those originating from Mae Sot in Thailand, Attapeu in Laos, or Ramu in Bangladesh (Fig. 1A), Supplementary Table 1). Gametocyte levels were also higher in samples from the Democratic Republic of the Congo compared to Bangladesh, despite both sites having only K13 wild-type isolates (Fig. 1A-B). These findings suggest that genetic background is an essential contributor to transmission potential. A considerable variation in the male to female gametocyte ratio across and within the different sites was observed, possibly reflecting variation observed during the course of an infection^14^ (Supplementary Fig. 1B and Supplementary Table 1). A significant positive but weaker association was also observed between asexual parasite density and the male to female gametocyte ratio (r=0.23, *p*=0.005; (Supplementary Fig. 1C). When we stratified the gametocytes and male to female gametocyte ratios by sampling site, we found in Pailin that infections with K13 mutants had slightly lower gametocyte gene expression levels than infections with wild-type parasites (∼1.3-fold, adjusted *p*=0.03; Fig. 2 A-B). However, the ratio of male to female gametocytes was significantly higher in K13 mutant than wild-type infections by ∼2-fold (adjusted *p*=0.004; Fig. 2C, Supplementary Table 1). This suggests that the Pailin K13^C580Y^ parasites may be more infectious to mosquitoes and support higher rates of outbreeding and transmission.

**Figure 1.**
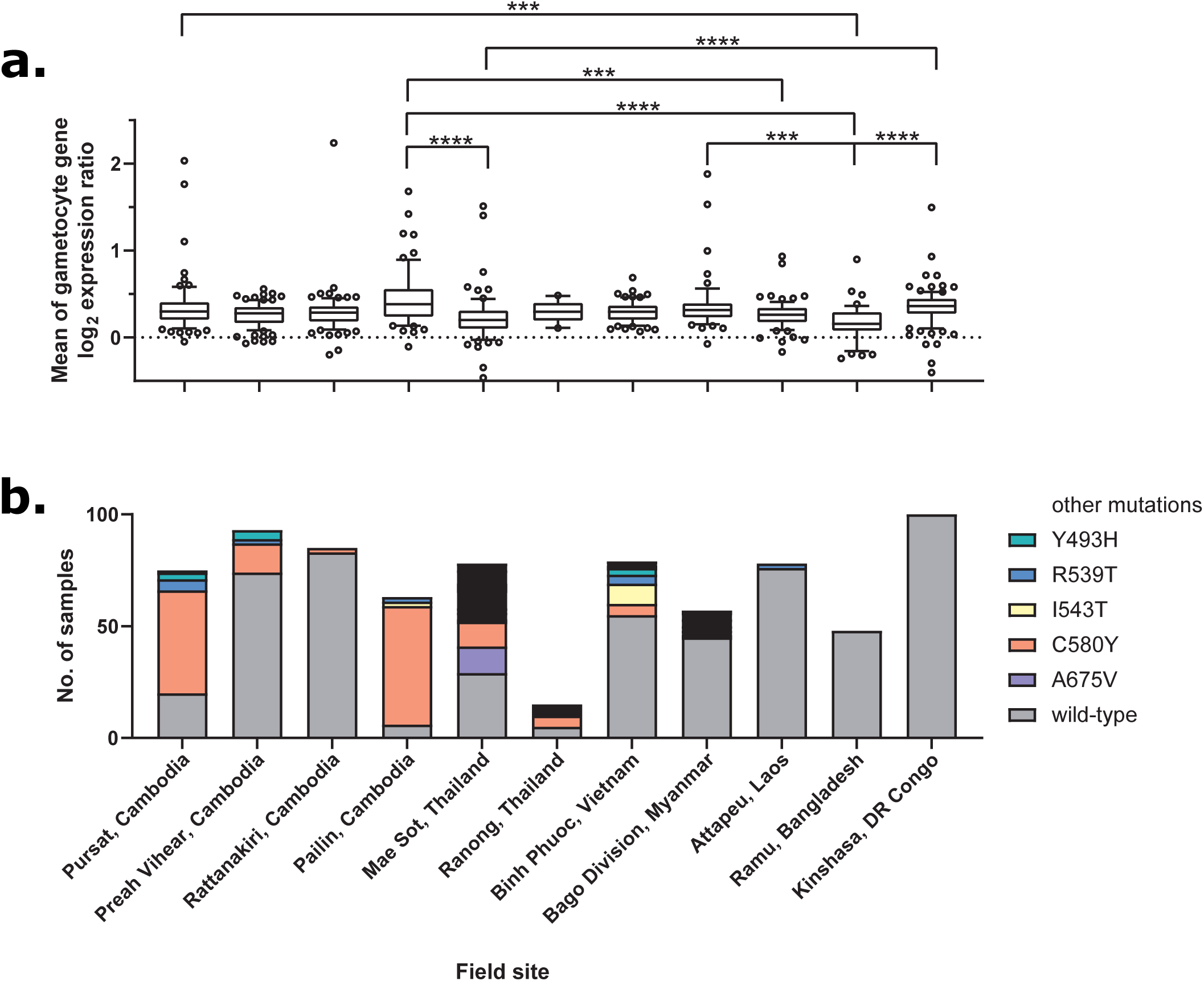
Levels of gametocyte transcripts in symptomatic clinical samples collected from the TRAC study in 2011-2013. Levels of gametocyte transcripts in symptomatic clinical samples collected from the TRAC study in 2011-2013. a) Box and whisker plots represent the median and 10-90 percentile of the mean of 121 gametocyte gene log_2_ expression ratios in 771 clinical isolates across 11 field sites. *** adjusted *p*<0.001, **** adjusted *p*<0.0001. Only statistically significant pairwise comparisons with adjusted *p*<0.001 as calculated by Dunn’s multiple comparisons test following Kruskal-Wallis test are shown. b) Proportion of K13 mutant genotypes among the 771 clinical samples.

**Figure 2.**
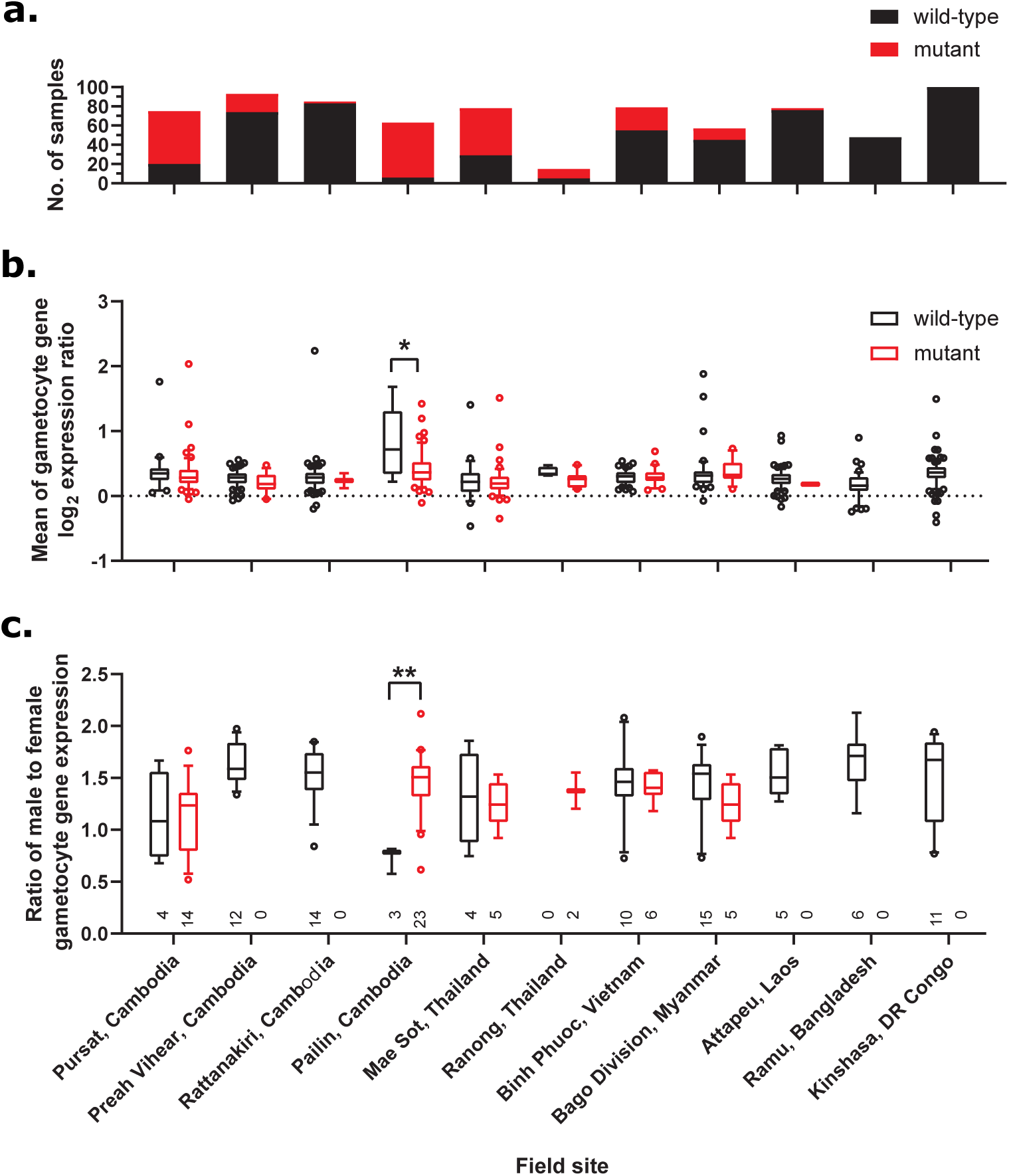
Distribution of gametocyte levels and male to female gametocyte ratios by K13 genotype across 11 field sites. a) Number of K13 wild-type or mutant samples in each field site (N=771). b) Box and whiskers represent the median and 10-90 percentile of gametocyte transcript levels stratified by K13 genotype status in each field site for 771 clinical isolates. This analysis suggested a slightly lower gametocyte prevalence for K13 mutants relative to wild-type isolates in Pailin, Cambodia. c) Box and whiskers represent the median and 10-90 percentile of male to female gametocyte ratios stratified by K13 genotype status (red – mutant; black - wild-type), in each site for 139 clinical samples (where the average male or female gametocyte transcript levels were higher than the asexual reference pool by ≥1.42-fold). Numbers listed are the number of clinical samples for K13 mutant or wild-type used in the analyses. Of the five field sites analyzed, the data showed a 2-fold higher proportion of male gametocyte in K13 mutants relative to wild-type isolates in Pailin, Cambodia. * adjusted *p*<0.05, ** adjusted *p*<0.01. See Supplementary Table 2 for transcript levels in isolates. As a point of reference, the ratio of male to female gametocytes for *in vitro* cultured 3D7 mature gametocytes profiled by the same microarray platform is 0.69 which is lower than the clinical samples.

### Effect of artesunate on the emergence of gametocytes in the K13 versus 3D7 study

Artesunate treatment initially reduced the total parasitaemia in all volunteers infected with either ART-resistant (n=13) or ART-sensitive parasites (3D7) (n=9) (Fig. 3A). ART-resistant infected volunteers experienced recrudescence earlier (Day 11) than ART-sensitive infected volunteers (between Day 15 and Day 20).

**Figure 3.**
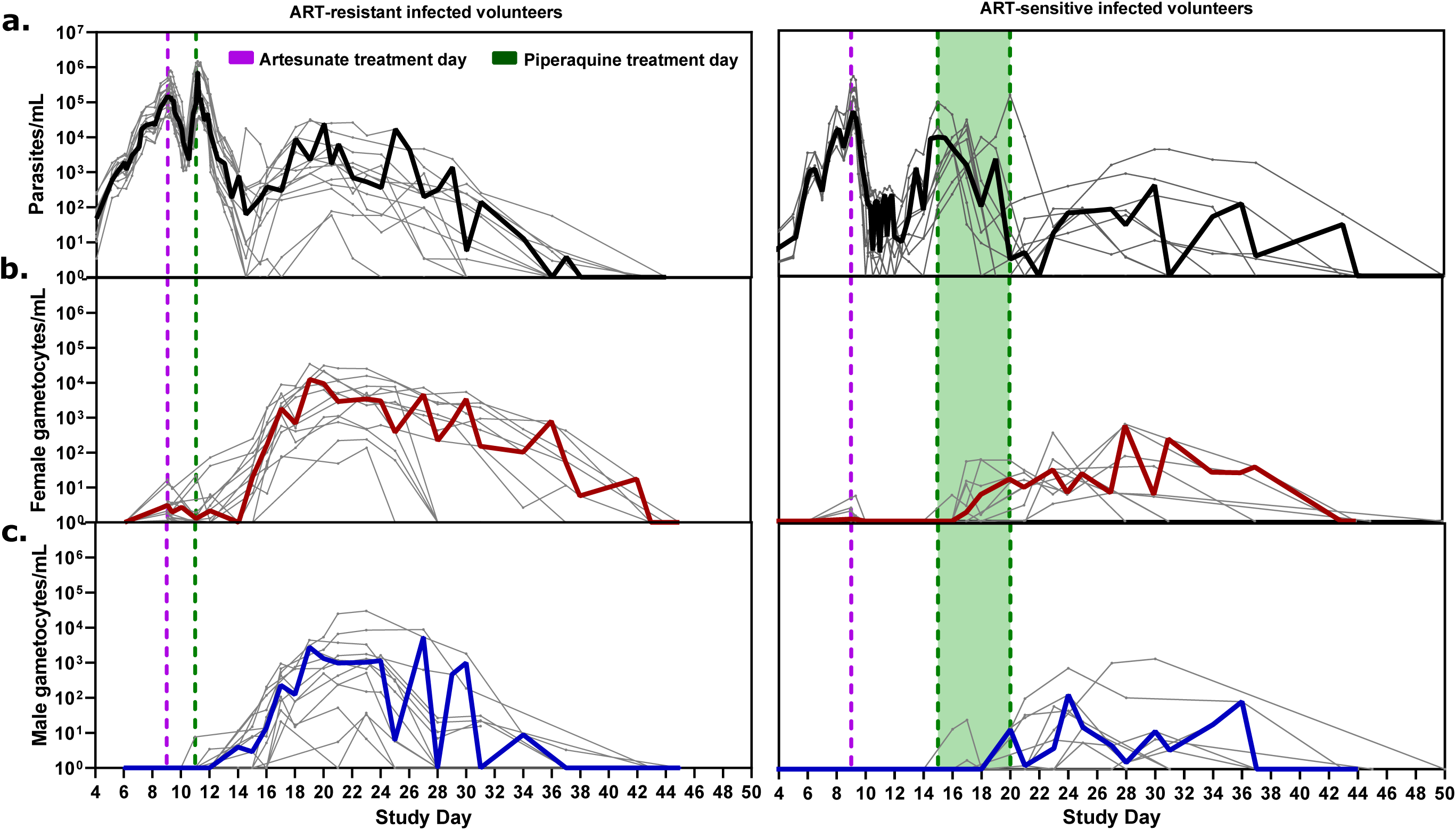
Parasitaemia counts before and after artesunate and piperaquine treatment. a) Total parasitemia, b) female, and c) male gametocytemia counts in volunteers infected with ART-resistant (left) and ART-sensitive (right) parasites. Purple and green lines indicate the time points when artesunate and piperaquine were administered, respectively. AP and primaquine treatment days are described in Supplementary Table 3. Thick coloured lines are the median values per day and thin grey lines are the individual values; non-detect data points were substituted by 1.

Accordingly, the peak of female and male gametocytemia occurred earlier among volunteers infected with ART-resistant parasites. Female gametocytemia peaked at a median of 12 days post-artesunate (range: 10-14 days) and male gametocytemia at 14 days post-artesunate (range: 10-18 days) in volunteers infected with ART-resistant parasites, while in volunteers infected with ART-sensitive parasites the female and male gametocytaemia both peaked at 17 days post-artesunate (female range:15-21 days and male range:14-21 days); *p*<0.001) (Figure 3B-C and Supplementary Fig. 3).

A positive correlation between total parasitaemia pre-artesunate and total gametocytemia 12 days post-artesunate was observed in volunteers infected with ART-resistant parasites (Supplementary Fig. 2B and Supplementary Table 2). In contrast, no correlation between total parasitaemia AUC pre-artesunate and total gametocytemia AUC 12 days post-artesunate was observed in volunteers infected with ART-sensitive parasites (Supplementary Fig. 2B and Supplementary Table 2). These data suggest, as previously reported, that artesunate inhibits the development of young gametocytes in ART-sensitive strains^18^.

The male to female gametocyte ratio varied over time in each volunteer (Supplementary Figure 4). We log_10_-transformed the ratio and calculated the mean in three periods: <20 days post-inoculation, 20-22 days post-inoculation and >22 days post-inoculation, and then compared between groups. Overall, the male to female ratio was higher among volunteers infected with ART-resistant than ART-sensitive parasites (0.200 vs 0.096, *p*=0.004; Supplementary Table 4), translating to ratios of 2:10 and 1:10 male to female gametocytes among volunteers with ART-resistant versus ART-sensitive infections respectively (Supplementary Table 4).

**Figure 4.**
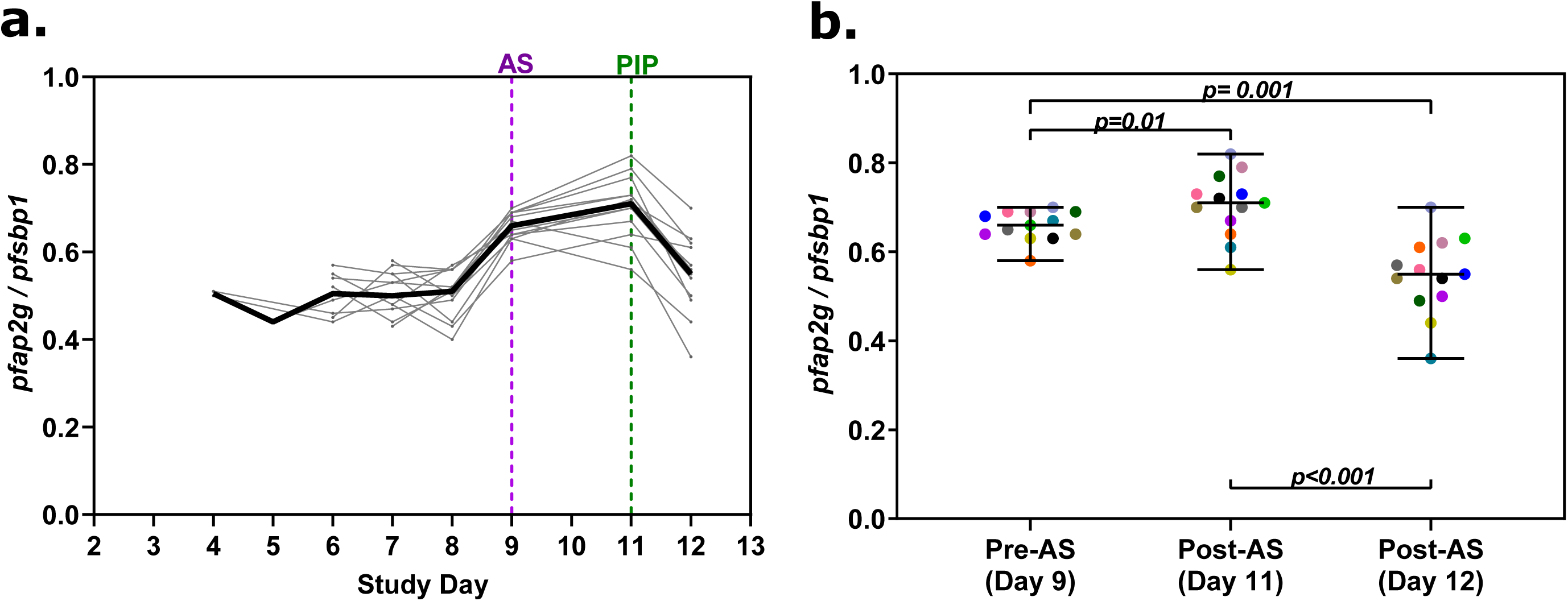
Sexual commitment rate increased after artesunate treatment in ART-resistant infected volunteers. a) Sexual commitment rate (ratio of log_10_ gametocyte ring transcript *pfap2g* / log_10_ ring transcripts *pfsbp1*) over time, as measured by qRT-PCR. The purple dashed line indicates administration of 2 mg/kg of artesunate; the green dashed line indicates administration of 960 mg/kg of piperaquine; the solid black line is the median of *pfap2g/pfsbp1* ratio per day; and grey lines are the *pfap2g/pfsbp1* ratio for each participant. b) Sexual commitment rate was significantly higher on Day 11 than on Day 9 or 12 (Wilcoxon-paired test). Colours indicate each of the 13 ART-resistant infected volunteers. AS: artesunate; PIP: piperaquine.

### Effect of artesunate on gametocyte production in volunteers infected with ART-resistant parasites

The *in vivo* sexual commitment rate did not differ significantly between volunteers infected with ART-resistant versus ART-sensitive parasites before artesunate treatment (Day 9), (median: 0.66, [range: 0.58-0,70; n=13] vs 0.63, [range: 0.55-0.67; n=3]). However, after artesunate treatment (Day 11) volunteers infected with ART-resistant parasites experienced recrudescence and had significantly higher sexual commitment rates during recrudescence after artesunate treatment (Day 11) compared to before treatment (Day 9) (Fig. 4A-B). In contrast, ART-sensitive parasite infections did not recrudesce by Day 11 and young gametocytes were not detected after artesunate treatment (Day 11) in volunteers infected with ART-sensitive parasites.

After 15 days of e*x vivo* culture, samples from volunteers infected with ART-resistant parasites taken after artesunate treatment (Day 11) yielded more mature male and female gametocytes than samples taken before artesunate treatment (Day 9) (*p*=0.02, Fig. 5A).This supports our findings that more young gametocytes were present post-artesunate treatment than pre-artesunate in the ART-resistant infected volunteers.

**Figure 5.**
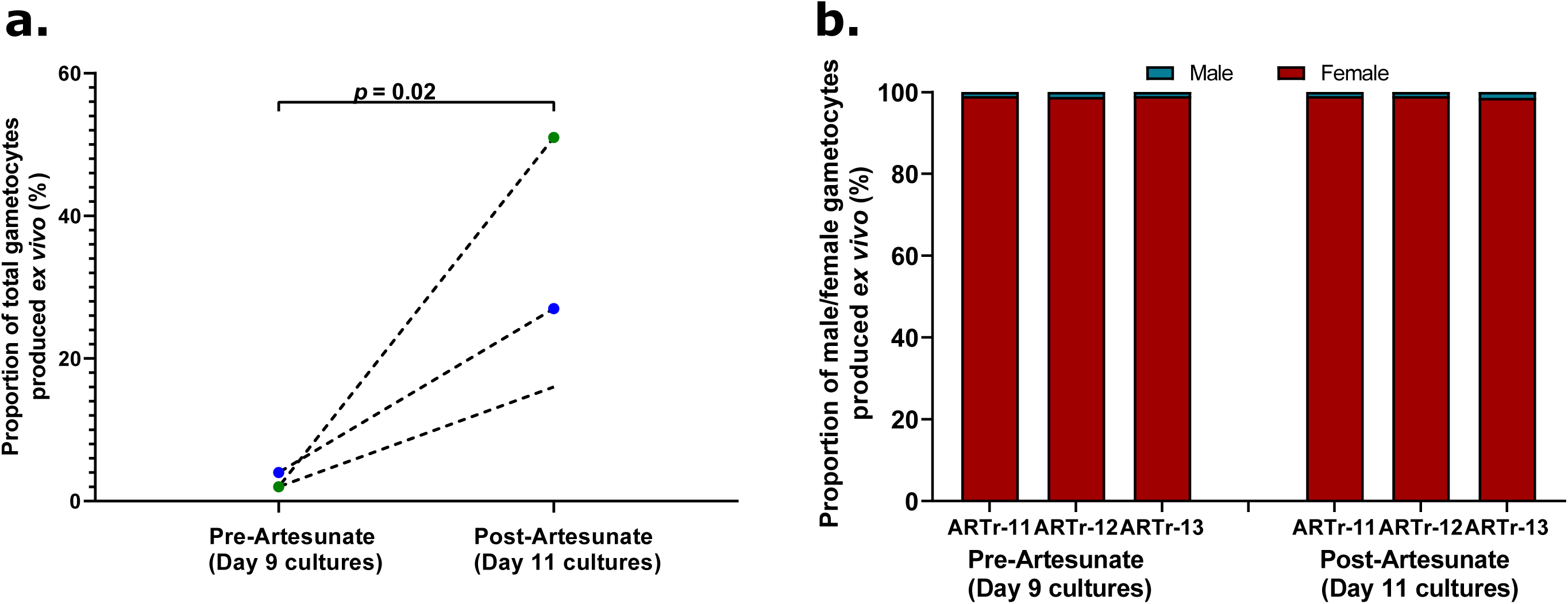
*Ex vivo* gametocyte cultures from post-artesunate samples developed more gametocytes than those from pre-artesunate samples, in volunteers infected with ART-resistant parasites. a) Proportion of total gametocytes (male and female) developed after 15 days of *ex vivo* culture as measured by qRT-PCR for *pfs25* and *pfMGET*, and normalized by total parasitaemia (18S rRNA), in samples taken pre and post artesunate treatment (ratio paired *t-*test); b) Bar-plots showing the sex composition of the *ex vivo* cultures 15 days after sample collection on Day 9 and Day 11.

Moreover, the proportion of male gametocytes was similar in parasite cultures taken pre- and post-artesunate treatment (pre-treatment: 0.9–1.2% vs post-treatment: 0.9– 1.4%) in volunteers infected with ART-resistant parasites (Fig. 5B), suggesting that artesunate did not affect male to female gametocyte ratios.

Before artesunate treatment (Day 9), blood samples taken from volunteers infected with ART-resistant and ART-sensitive parasites yielded a relatively similar number of gametocytes (ARTr: 2.1%, 2.2% and 3.9%, Fig 5A) vs (ARTs: 0.5%, 2.7% and 4.2%, Supplementary Fig 5A), and with a similar male to female gametocyte ratio (Fig 5B and Supplementary Fig 5B). However, samples taken from volunteers infected with ART-sensitive parasites after artesunate treatment (Day 11) did not develop gametocytes, consistent with our data showing a loss of viable rings on Day 11 following treatment. (Supplementary Fig. 5A).

### Effect of AP on the infectiousness to *Anopheles* mosquitoes of volunteers with ART-resistant parasite infection

Transmissibility of volunteers to *An. stephensi* mosquitoes was analysed by DFA and eMFA in 12/13 volunteers infected with ART-resistant parasites (Supplementary Fig. 6).

**Figure 6.**
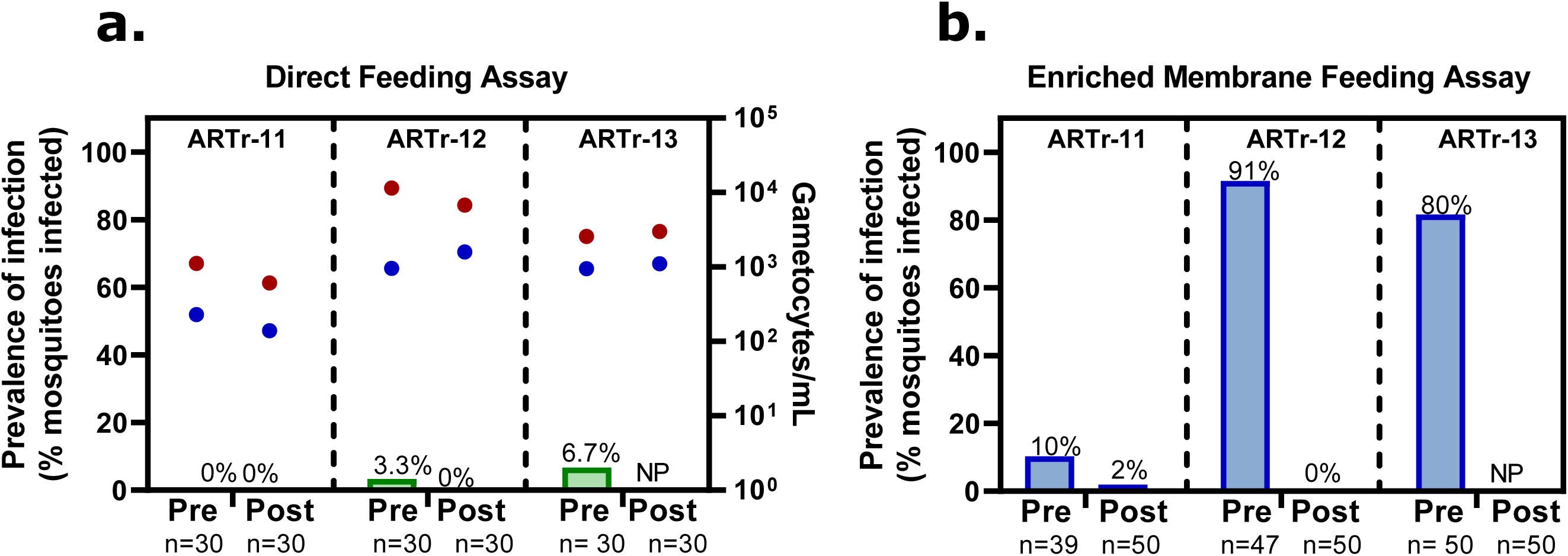
Prevalence of mosquito infection pre and post-atovaquone-proguanil (AP) treatment on Day 21 in ART-resistant (ARTr) volunteers. a) Infections were carried out by Direct Feeding Assay (DFA) and green bars indicate the proportion of infected midguts, red circles indicate female gametocytemia (gametocytes/mL), and blue circles indicate male gametocytemia (gametocytes/mL) detected by qRT-PCR at the time of feeding, n= number of mosquitoes used in each assay b) Infections were carried out by enriched Membrane Feeding Assay (eMFA) and blue bars indicate the proportion of infected midguts, n= number of mosquitoes used per assays AP. NP: Not performed.

Among volunteers that did not receive AP treatment prior to conducting the transmission assays (n=7), four were infectious to *An. stephensi* mosquitoes by DFA (57%), and six by eMFA (86%). The median mosquito infection rate among volunteers infected with ART-resistant parasites by DFA was 5% (range: 3–10%, n = 4) and by eMFA was 74% (range: 20–97%; n = 6) (Supplementary Fig. 7A). The four successful infections by DFA had significantly higher male (Mann-Whitney *U* test; *p*= 0.03) and female (Mann-Whitney *U* test; *p*= 0.03) gametocyte levels than those that were not infectious on the day when transmission assays were performed (Supplementary Fig. 7B).

**Figure 7.**
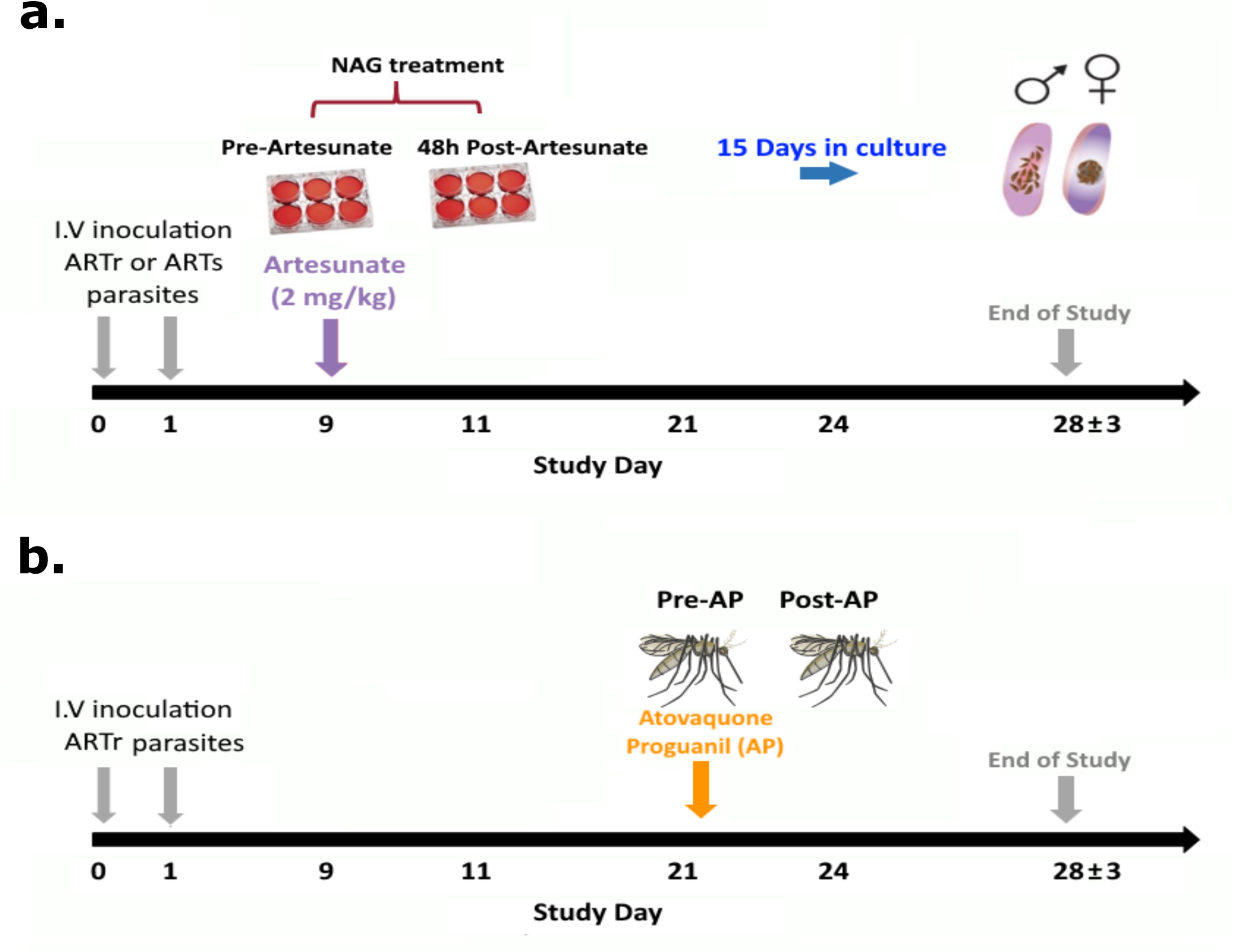
Study designs using the ART-resistant IBSM mode. a) *ex vivo* gametocyte production in ART-resistant (ARTr) and ART-sensitive (ARTs) infected volunteers. b) effect of atovaquone-proguanil (AP) on transmission in three ART-resistant volunteers enrolled in the K13 versus 3D7 clinical trial.

Among volunteers that received AP treatment prior to conducting the transmission assays (n=5), only two were infectious to mosquitoes, one by DFA with an infection rate of 4% and one by eMFA (Supplementary Fig. 8), with an infection rate of 1.3%. Interestingly, these five volunteers also had the highest levels of male and female gametocytes among the volunteers infected with ART-resistant parasites at the time of the transmission assays (Supplementary Fig. 8). Thus, in the last cohort of the trial, we decided to evaluate the effect of AP on transmission by conducting a paired comparison, pre- and post-AP treatment with three volunteers infected with ART-resistant parasites. The mosquito infection rates for the three volunteers pre-AP treatment were 0%, 3% and 6.7% (n = 30 mosquitoes) by DFA (Fig. 6A) and 10% (n = 39 mosquitoes), 80% (n=50 mosquitoes) and 91% (n = 47 mosquitoes) by eMFA (Fig. 6B). The infection rates for two volunteers post-AP treatment were all negative by DFA (Fig. 6A), and only one was positive 2% (n= 50 mosquitoes) by eMFA (Fig. 6B). No significant difference in gametocytemia was found between pre- and post-AP among ART-resistant infected volunteers (female: pre-AP median: 2,569 [range: 1,115 – 11,449] vs post-AP median: 3,449 [range: 475-3,551] gams/mL; *p*= 0.58; male: pre-AP median 978 [range: 236-987] vs post-AP median: 670 [range: 150-1282] gams/mL; *p=*0.67) (Fig. 6A).

## DISCUSSION

In this study, we investigated the transmissibility of parasites carrying K13 mutations, with the underlying hypothesis that increased transmissibility may be a factor in the spread of ART-resistant strains across the Greater Mekong Subregion. Using an experimental infection model with Cam3II^R539T^ parasites carrying the K13 mutant allele, we showed that a single dose of artesunate increased the sexual commitment rate in volunteers infected with these parasites compared to volunteers infected with the ART-sensitive reference strain 3D7.

Moreover, a survey of male and female gametocyte transcriptomes obtained from patients from various geographical regions revealed a higher ratio of male to female transcriptional signatures particularly in K13 mutants from Pailin, Cambodia, where artemisinin resistance is well established. Although higher overall gametocyte levels have been shown to contribute to increased transmissibility to *Anopheles* mosquitoes^19^, mathematical modelling has also predicted that a high male to female gametocyte ratio in low-density infections could enhance the chances of successful transmission^20,21^. Thus, the higher male to female gametocyte ratio among K13-mutant infections compared to K13 wild-type in Pailin, a hot-spot of ART resistance, suggests that this could be a contributor to the spread of resistance in the region.

We also assessed, for the first time, the *in vivo* effect of artesunate on the sexual commitment rate of ART-resistant and ART-sensitive parasites. In this study, a single dose of artesunate cleared ART-sensitive asexual parasitemia to a level where young gametocytes could no longer be detected but enhanced the sexual commitment rate in infections with the ART-resistant parasite. This is a key transmission advantage in areas where ART resistance is not present and therefore transmission-blocking antimalarials, such as primaquine, are not routinely administered^22^. Similarly, an increase on gametocytemia, after a 6-hour DHA pulse *in vitro* assay with 700 nM DHA, has been reported in a *P. falciparum* strain with a different *k13* allele conferring artemisinin resistance (*Pf* IPC-5202^C580Y^)^11^. Although *in vitro* assays measuring sexual commitment rates have been described, the lengthy protocols make them impractical for large field studies^23,24^. Thus, quantification of the dynamic changes in the sexual commitment rate of parasites in field studies has been scarce. However, a new approach combining *ex vivo* culture and molecular assays targeting young gametocytes such as *pfap2g* and *pfgdv1* proved useful in characterising sexual commitment rates in isolates from Ghana^25^. A field study characterising sexual commitment rates in areas of ART resistance would contribute to our understanding of the overall transmission potential of these strains.

Likewise, considering the evidence on sexual commitment plasticity in a murine *P. chabaudi* parasite model in response to environmental factors (e.g. drop in haematocrit, drug pressure)^26^ and the results presented herein, further investigation on the effect of current antimalarial regimes on gametocyte production in ART-resistant strains is necessary.

Although we did not investigate the mechanism by which artesunate enhanced the sexual commitment rate in ART-resistant infections, a number of possible mechanisms would be worth investigating in future studies. For instance, assessing whether artesunate treatment results in changes in host-factors such as lysophosphatidylcholine (LysoPC) content in the plasma^27^, thereby modulating sexual commitment in the surviving parasites. Another possible explanation is that the ART resistance mechanisms of these strains may have a direct or indirect influence on the sexual commitment rate of drug-resistant parasites. ART-resistant parasites undergo metabolic changes resulting in reduced haemoglobin endocytosis^28^. It is possible that the lack of amino acids could generate a general cellular stress response in the parasite^29,30^. Thus, gametocytogenesis might be triggered as a collateral effect of the perceived starvation state in the parasite^26^.

This study used the ART-sensitive strain 3D7. Ideally, the comparator strain for the investigations performed herein should be an isogenic parasite such as the reverted K13 wild-type Cambodian strain (Cam3.II^rev^) with an identical genetic background as the K13 mutant, Cam3.II^R539T^ used in this study. Unfortunately, logistical difficulties inherent in producing a clinical-grade bank of such parasites enabling a clinical trial prevented us from including this control experiment. Hence, we cannot exclude the possibility that other genetic differences between the Cam3.II^R539T^ parasite and the reference strain 3D7 may contribute to the observed effect in sexual commitment rates. These results may be explained by *in vitro* data, where other non-K13 polymorphisms modulate sexual commitment rates and thus may explain the observed higher level of gametocytemia in areas with slower parasite clearance after treatment^1,6^. More research is required to fill the gaps in identifying determinants of transmission of ART-resistant parasites in endemic settings, including the genetic background of the parasites.

Using a combination of molecular tools and *ex vivo* culture, we found that artesunate treatment did not substantially affect the proportion of male gametocytes in ART-resistant infections. These results support those of Witmer et al.^9^, whereby ART-resistant male gametocytes carrying the *k13* R539T and C580Y mutations from Southeast Asia were not affected by artemisinin derivatives. This suggests that under artemisinin drug pressure, an ART-resistant strain that is a naturally higher male gametocyte producer will continue to transmit better than a sensitive strain, which is a significant concern considering the observed male gametocyte bias seen in ART-resistant K13 mutant isolates from Pailin, Cambodia.

To prevent the spread of ART-resistant parasites in areas where they are prevalent, the WHO recommends giving a single low-dose of primaquine (SLD PQ, 0.25mg/kg or 15mg), the only widely available antimalarial that targets mature gametocytes, added to a standard antimalarial treatment^31^. However, new antimalarials with transmission-blocking effect are needed as SLD PQ can still cause haemoglobin loss in G6PD deficient patients^22^. We showed that a 3-day course of AP was sufficient to prevent the infection of *Anopheles* mosquitoes. These results support further assessment of atovaquone or AP as transmission-blocking antimalarial in areas where ART resistance is prevalent. In support of these data, atovaquone was recently shown to synergize with dihydroartemisinin against Cam3.II^R539T^ parasites, while not showing this synergy against Cam3.II^WT^ parasites (Mok *et al*., manuscript under review).

The transmission-blocking effect of AP has been established *in vitro* with an artemisinin-sensitive strain in standard MFAs^38^. Atovaquone was also effective at blocking *P. falciparum* transmission when applied ectopically to infected *Anopheles* mosquitoes^39^. However, this is the first time that the transmission-blocking effect of AP has been tested in an experimental human infection model with an ART-resistant parasite. Although there has been a reluctance to use this drug combination, due to the rapid development of atovaquone resistance in the parasite^32,33^, evidence showing that such mutations preclude onward transmission of resistant parasites in the mosquito^34^ suggests further consideration of this approach.

We note that in this study the low number of transmission events observed in the clinical trial did not allow us to determine whether enhanced sexual commitment rates after drug exposure in volunteers infected with ART-resistant parasites translated into increased infectivity overall. However, several studies have reported enhanced infectivity in resistant parasites after treatment^35-38^. Other uncontrolled variables including gametocyte maturity^39,40^, human host genetic factors^41-44^, and immune responses^45,46^, could significantly influence the transmission potential of a *P. falciparum* infection.

In conclusion, this study suggests that ART-resistant infections have a greater transmission potential than ART-sensitive strains. Thus, these data have implications in areas where ART resistance is emerging, as it shows how sub-curative doses of an artemisinin component can favour the propagation of isolates with resistant phenotypes. Moreover, it highlights the importance of considering ART-resistant parasites from various geographical backgrounds when developing antimalarials that target *P. falciparum* sexual stages, as their effectiveness can vary based on the parasite’s genetic makeup. Finally, this study suggests an additional benefit of a 3-dose AP treatment as a transmission-blocking antimalarial in areas where ART resistance is prevalent.

## METHODS

### Clinical trials

#### Tracking Resistance to Artemisinin Collaboration I (TRAC I) study

Transcriptome data previously generated for the clinical malaria samples that were collected under the multi-collaborative TRAC I study between 2011 and 2013 were used in this analysis^1,29^. The TRAC I microarray data is accessible through GEO Series accession number GSE59099 (http://www.ncbi.nlm.nih.gov/geo/query/acc.cgi?acc=GSE59099). At the time of collection, ART resistance was either established (in Pursat and Pailin, Cambodia; Mae Sot and Ranong, Thailand; and Binh Phuoc, Vietnam), newly emerging (in Preah Vihear and Rattanakiri, Cambodia; Attapeu, Laos; and Shwe Kyin, Myanmar) or absent (in Ramu, Bangladesh; and Kinshasa, Democratic Republic of Congo (DR Congo)) ^1^. Sites with fewer than 15 isolates were excluded (Sisakhet and Khun Han in Thailand; and Ilorin in Nigeria). We also removed isolates that showed heterozygosity or missing genotype data in the *k13* allele^1^. In total, we selected 771 out of 1048 clinical samples from 10 sampling sites in Southeast Asia and one in Africa.

Briefly, whole blood was collected from patients arriving at health facilities upon admission. After white blood cell depletion using CF11 columns, RNA was extracted using TRIzol®^29^. cDNA was synthesized and amplified using a 5’-3’ reverse-transcriptase template switching-PCR protocol (SMART) and then hybridized against a reference pool consisting of 3D7 asexual blood stages on a custom long-oligonucleotide microarray chip as described previously^29^. The average log_2_ expression ratios of 121 highly abundant gametocyte stage IV and V genes were used as an index to quantify gametocytes present in the samples^47^. The male to female gametocyte ratio was estimated by calculating the ratio of the average transcript abundance of six male-specific gene markers to the average transcript abundance of 14 female-specific gene markers (listed in Supplementary Table 5). To ensure that this analyses was based on samples with either female or male gametocytes present, we only considered samples that showed average transcript abundance in either male or female gametocyte genes of at least 1.42-fold higher than the asexual reference pool (log_2_ expression >0.5) (Supplementary Table 1). This totalled 139 samples.

This study was approved by the relevant local ethics committees and the Oxford Tropical Research Ethics Committee. All adult patients or the parents of children gave written informed consent.

#### The ART-resistant IBSM model

An IBSM volunteer infection study was undertaken^17^ to compare parasite clearance after single-dose artesunate in volunteers infected with either ART-resistant Cam3.II^R539T^ parasites or the ART-sensitive reference strain 3D7^17^. This clinical trial, referred to herein as the K13 vs 3D7 trial, was a phase 1, open-label trial with 3 cohorts, conducted at Q-Pharm Pty Ltd, Brisbane, Australia^17^.

Thirteen healthy malaria-naïve volunteers were inoculated with ART-resistant parasites (Cam3.II^R539T^) on Day 0, and nine volunteers with 3D7 ART-sensitive parasites on Day 1. Parasitaemia was monitored by qPCR of the 18S-rRNA gene^48^ from Day 4 post-inoculation. A single dose of artesunate (2 mg/kg; Guilin Pharmaceutical Shanghai Co. Ltd) was given to all infected volunteers on Day 9 to treat asexual parasitaemia. Piperaquine (960 mg, PCI Pharma Services) was given to all ART-resistant infected volunteers on Day 11 to treat the first recrudescence, whereas for ART-sensitive infected volunteers piperaquine was given between Day 15 and Day 20 when asexual parasitemia recrudesced (Supplementary Table 3). AP (Malarone^®^, GlaxoSmithKline Australia) was given between Day 16 and Day 28 to treat the second recrudescence or as a compulsory treatment at the end of the study for all volunteers (Supplementary Table 3). Primaquine was given between Day 21 and Day 35 to eliminate gametocytes at the end of the study (Supplementary Table 3).

Mature female and male gametocyte levels were measured from Day 9 by quantitative reverse transcriptase PCR (qRT-PCR) of transcripts *pfs25* and *pfMGET* (for female and male gametocytes, respectively, and were converted to gametocytes/mL as previously described^49^.

The K13 versus 3D7 trial was approved by the QIMR Berghofer and Australian Red Cross Blood Service Human Research Ethics Committees. All volunteers gave written informed consent before enrolment. The trial was registered with the Australian New Zealand Clinical Trials Registry: ACTRN12617001394336.

#### Gametocyte dynamics after artesunate treatment in the K13 vs 3D7 trial

The area under the curve (AUC) of total parasitaemia and total gametocytemia (sum of female and male gametocytes) was calculated to determine the magnitude of the correlation between them^14^. The AUC of the total parasite density was calculated using consecutive parasitaemia/mL data from Day 4 until the day of artesunate treatment (Day 9), and the AUC of the total gametocytemia^14,49^ was from the first positive sample (male and female) until 12 days post artesunate administration (Day 21). This analysis assumed 12 days as the period it would take for gametocytes to mature and return to circulation.

The male to female gametocyte ratio was determined by dividing the number of male gametocytes over female gametocytes/mL. For accurate quantification, we only included time points when both male and female gametocytes were greater than 6 gametocytes/mL, which is the lower limit of the reportable range for both gametocyte assays^49^.

#### Effect of artesunate on sexual commitment rate

We evaluated the effect of artesunate on the proportion of young gametocytes produced per asexual cycle, i.e. the sexual commitment rate. This analysis used a qRT-PCR assay targeting the *pf*AP2 transcription factor (*pfap2-g, PF3D7_1466400*) that is expressed in young gametocyte stages and is required for gametocytogenesis^25^, as well as the ring-stage transcript skeleton-binding protein 1 (*pfsbp1, PfE0065w*) expressed by asexual ring-stages and young gametocytes^14,50^. The sexual commitment rate was defined as the ratio between the log_10_-transformed levels of *pfap2* and *pfsbp1* transcripts, as measured in samples collected from Day 4 until Day 12. These time points included the window when young gametocytes stages induced during artesunate treatment first appeared in the blood circulation (i.e. approximately 48 hours after drug administration)^51^.

An *ex vivo* approach was used to determine whether artesunate induces a change in the male to female gametocyte ratio. Briefly, blood samples were taken pre-(Day 9) and post-artesunate treatment (Day 11) from volunteers infected with ART-resistant and ART-sensitive parasites. Asexual parasites were removed with 50 mM of N-Acetyl Glucosamine, (NAG, Sigma-Aldrich), and young gametocytes present in the samples were allowed to mature to stage V under standard *in vitro* culture conditions (5% haematocrit, RPMI complete media (Sigma-Aldrich), 10% human serum, and a fixed gas composition (5% O_2_, 5% CO_2_, 90% N_2_)^52^. After 15 days, total parasitemia was measured in the culture pellets by qRT-PCR specific to the 18S-rRNA gene, and gametocyte density was measured by qRT-PCR targeting *pfs25* and *pfMGET* transcripts^49^ (Fig. 7A).

#### Effect of AP on the transmissibility of ART-resistant infected volunteers to *Anopheles* mosquitoes

Transmissibility of gametocytes from volunteers infected with ART-resistant parasites was assessed by direct skin feeding assays (DFA) and membrane feeding assays using gametocytes enriched from large blood volumes via Percoll gradient centrifugation (eMFA)^14^. DFA and eMFA were performed between Day 21 and Day 24, on consenting volunteers presenting with more than 100 female gametocytes/mL on the day of these assays.

For DFAs, volunteers were exposed to approximately 30 starved female *Anopheles stephensi* mosquitoes (Sind-Kasur Nijmegen strain) that were allowed to feed for 15 minutes on the forearm. For eMFA, parasite-infected erythrocytes were enriched from 36 mL of whole blood using a 65% Percoll gradient. The gametocyte-enriched layer was mixed with uninfected red blood cells and AB serum (from a malaria-naïve donor, Australian Red Cross Blood Service) and fed to 60–100 starved mosquitoes for 30 minutes using a membrane-feeding apparatus. Blood-fed mosquitoes were returned to controlled environmental conditions (30°C and 70-80% relative humidity with a 12-hour day/night cycle) and maintained on a sucrose-PABA solution for eight days^14^. *P. falciparum* infection in the mosquito midguts was determined by qPCR targeting the 18S-rRNA gene^53^. The mosquito infection rate was defined as the proportion of mosquitoes with infected midguts.

The effect of AP on transmission was investigated in three volunteers by comparing the gametocyte levels and mosquito infection rates by DFA and eMFA, before and after AP treatment (Fig. 7B).

### Statistical Methods

In the TRAC study, differences in gametocyte gene expression levels between field sites were assessed using Kruskal-Wallis and corrected for multiple comparisons using Dunn’s test. Differences between male to female ratios between the K13 mutant and wild-type samples for each field site was determined by *t*-test with correction for multiple comparisons using the Holm-Sidak method.

For the K13 vs 3D7 study, GraphPad Prism (version 7), was used for linear regression and to compute AUC measures. The pre-treatment parasitaemia AUC and corresponding total gametocytemia AUC were log_10_-transformed to assess their correlation. Mann-Whitney *U* tests were used to i) compare the AUC of total gametocytemia, from first positive until last positive, between ART-resistant and ART-sensitive infected volunteers; and ii) compare the day when male or female peak gametocytemia was reached between ART-resistant and ART-sensitive infected volunteers. R version 3.5.2 was used to calculate the male to female gametocyte ratio. Log_10_-transformed data and one-way ANOVA tests were used to examine the relationship between male to female gametocyte ratios and day groups. *t*-tests were used to compare male to female gametocyte ratios between ART-resistant and ART-sensitive infections in volunteers. The Wilcoxon’s matched-pairs signed-rank test was used to compare the sexual commitment rate before and after artesunate treatment, as well as gametocyte levels before and after AP treatment. The proportion of female and male gametocytes in culture was compared using a ratio paired *t test*. Finally, female and male gametocyte levels between infectious and non-infectious volunteers were compared using the Mann-Whitney *U* test.

### DATA AVAILABILITY

The authors declare that all other data supporting the findings of this study are available within the article and its supplementary Information files or are available from the authors upon request.

## ACKNOWLEDGEMENTS

The authors thank all volunteers who participated in the study, all staff from the clinical study site Q-Pharm Pty. Ltd and all staff from the QPID Laboratory for PCR analysis. From the QIMR Berghofer Medical Research Institute we thank Matt Adams and Melanie Rampton for technical assistance with mosquito rearing, Katharine Trenholme for assistance with inoculum preparation, Peter O’Rourke for statistical advice, and Laura Cascales for assistance with manuscript writing and editing support. From the University of Melbourne, we thank Pengxing Cao for critical review of the manuscript. From Uniformed Services University of the Health Sciences, Surendra K. Prajapati and Kim Williamson for sharing the protocol to determine sexual commitment rate. This study was supported by funds from Medicines for Malaria Venture (OPP1111147). JSM was supported by an Australian Government National Health and Medical Research Council (NHMRC) Practitioner Fellowship (APP1135955). JSM, MR and ZP were supported by an NHMRC Program Grant (1132975). DAF gratefully acknowledges funding from the NIH (R01 AI109023) and the US Department of Defence (to DAF and SM, W81-XWH-19-10-086). SM is the recipient of a long-term fellowship from the Human Frontiers of Science Program (LT000976/2016-L). We are extremely grateful to the TRAC clinical site coordinators, doctors, nurses, laboratory technicians, and patients who participated in the multi-site TRAC clinical study. TRAC was funded by the UK Department for International Development (DFID) for the benefit of developing countries and was coordinated by MORU Tropical Health Network which receives core funding from the Wellcome Trust. Whole-genome sequencing and genotyping for TRAC was funded by the Wellcome Trust through core funding of the Wellcome Trust Sanger Institute (098051).

## AUTHOR CONTRIBUTION

S.M. and D.A.F. conducted the transcriptomic analysis; K.A.C., R.E.W. J.J.M. and J.S.M. designed the K13 vs 3D7 clinical trial; G.J.R. and C.Y.T.W. performed molecular analysis in the K13 vs 3D7 clinical trial.; Z.P., H.M., S.L. and G.J.R. performed mosquitoes assays; J.G., M.R. and Z.P. conducted culturing experiments; Z.P., L.W., S.M. and L.M. performed formal statistical analysis; A.O., B.B. and J.S.M. conducted clinical assessment of volunteers; and J.S.M., M.W.A.D. and Z.P. designed the transmission experiments. Z.P., S.M., J.S.M. and D.A.F. wrote the manuscript, and all authors read the manuscript and approved its submission.

## REFERENCES

1 Ashley, E. A. et al. Spread of artemisinin resistance in Plasmodium falciparum malaria. N Engl J Med 371, 411–423, doi:10.1056/NEJMoa1314981 (2014).

2 Dondorp, A. M. et al. Artemisinin resistance in Plasmodium falciparum malaria. N Engl J Med 361, 455–467, doi:10.1056/NEJMoa0808859 (2009).

3 Mathieu, L. C. et al. Local emergence in Amazonia of Plasmodium falciparum k13 C580Y mutants associated with in vitro artemisinin resistance. Elife 9, doi:10.7554/eLife.51015 (2020).

4 Miotto, O. et al. Emergence of artemisinin-resistant Plasmodium falciparum with kelch13 C580Y mutations on the island of New Guinea. bioRxiv, 621813 (2019).

5 Uwimana, A. et al. Emergence and clonal expansion of in vitro artemisinin-resistant Plasmodium falciparum kelch13 R561H mutant parasites in Rwanda. Nat Med, doi:10.1038/s41591-020-1005-2 (2020).

6 Eksi, S. et al. Plasmodium falciparum gametocyte development 1 (Pfgdv1) and gametocytogenesis early gene identification and commitment to sexual development. PLoS Pathog 8, e1002964, doi:10.1371/journal.ppat.1002964 (2012).

7 Barry, A. et al. Increased gametocyte production and mosquito infectivity in chronic versus incident Plasmodium falciparum infections. medRxiv (2020).

8 Lozano, S. et al. Gametocytes from K13 propeller mutant Plasmodium falciparum clinical isolates demonstrate reduced susceptibility to dihydroartemisinin in the male gamete exflagellation inhibition assay. Antimicrob Agents Chemother 62, doi:10.1128/AAC.01426-18 (2018).

9 Witmer, K. et al. Artemisinin-resistant malaria parasites show enhanced transmission to mosquitoes under drug pressure. bioRxiv, 2020.2002.2004.933572, doi:10.1101/2020.02.04.933572 (2020).

10 Buckling, A., Ranford-Cartwright, L. C., Miles, A. & Read, A. F. Chloroquine increases Plasmodium falciparum gametocytogenesis in vitro. Parasitology 118 (Pt 4, 339-346, doi:10.1017/s0031182099003960 (1999).

11 Rajapandi, T. Upregulation of gametocytogenesis in anti-malarial drug-resistant Plasmodium falciparum. J Parasit Dis 43, 458–463, doi:10.1007/s12639-019-01110-w (2019).

12 Koepfli, C. & Yan, G. Plasmodium gametocytes in field studies: do we measure commitment to transmission or detectability?Trends Parasitol 34, 378–387, doi:10.1016/j.pt.2018.02.009 (2018).

13 Alkema, M. et al. A randomized clinical trial to compare P. falciparum gametocytaemia and infectivity following blood-stage or mosquito bite induced controlled malaria infection. J Infect Dis 222, 1416–1416, doi:10.1093/infdis/jiaa157 (2020).

14 Collins, K. A. et al. A controlled human malaria infection model enabling evaluation of transmission-blocking interventions. J Clin Invest 128, 1551–1562, doi:10.1172/JCI98012 (2018).

15 Reuling, I. J. et al. A randomized feasibility trial comparing four antimalarial drug regimens to induce Plasmodium falciparum gametocytemia in the controlled human malaria infection model. Elife 7, doi:10.7554/eLife.31549 (2018).

16 Beshir, K. B. et al. Residual Plasmodium falciparum parasitemia in Kenyan children after artemisinin-combination therapy is associated with increased transmission to mosquitoes and parasite recurrence. J Infect Dis 208, 2017–2024, doi:10.1093/infdis/jit431 (2013).

17 Watts, R. E. et al. Safety and parasite clearance of artemisinin resistant Plasmodium falciparum infection: A pilot and a randomised volunteer infection study in Australia. PLoS Med 17, doi:https://doi.org/10.1371/journal.pmed.1003203 (2020).

18 Price, R. N. et al. Effects of artemisinin derivatives on malaria transmissibility. Lancet 347, 1654–1658, doi:10.1016/s0140-6736(96)91488-9 (1996).

19 Churcher, T. S. et al. Predicting mosquito infection from Plasmodium falciparum gametocyte density and estimating the reservoir of infection. Elife 2, e00626, doi:10.7554/eLife.00626 (2013).

20 Bradley, J. et al. Predicting the likelihood and intensity of mosquito infection from sex specific Plasmodium falciparum gametocyte density. Elife 7, doi:10.7554/eLife.34463 (2018).

21 Paul, R. E., Brey, P. T. & Robert, V. Plasmodium sex determination and transmission to mosquitoes. Trends Parasitol 18, 32–38, doi:10.1016/s1471-4922(01)02122-5 (2002).

22 Bancone, G. et al. Single low dose primaquine (0.25 mg/kg) does not cause clinically significant haemolysis in G6PD deficient subjects. PloS One 11, e0151898, doi:10.1371/journal.pone.0151898 (2016).

23 Brancucci, N. M., Goldowitz, I., Buchholz, K., Werling, K. & Marti, M. An assay to probe Plasmodium falciparum growth, transmission stage formation and early gametocyte development. Nat Protoc 10, 1131–1142, doi:10.1038/nprot.2015.072 (2015).

24 Lucantoni, L., Duffy, S., Adjalley, S. H., Fidock, D. A. & Avery, V. M. Identification of MMV malaria box inhibitors of Plasmodium falciparum early-stage gametocytes using a luciferase-based high-throughput assay. Antimicrob Agents Chemother 57, 6050–6062, doi:10.1128/AAC.00870-13 (2013).

25 Usui, M. et al. Publisher Correction: Plasmodium falciparum sexual differentiation in malaria patients is associated with host factors and GDV1-dependent genes. Nature Commun 10, 2740, doi:10.1038/s41467-019-10805-w (2019).

26 Schneider, P. et al. Adaptive plasticity in the gametocyte conversion rate of malaria parasites. PLoS Pathog 14, e1007371, doi:10.1371/journal.ppat.1007371 (2018).

27 Brancucci, N. M. B. et al. Lysophosphatidylcholine regulates sexual stage differentiation in the human malaria parasite Plasmodium falciparum. Cell 171, 1532–1544 e1515, doi:10.1016/j.cell.2017.10.020 (2017).

28 Birnbaum, J. et al. A Kelch13-defined endocytosis pathway mediates artemisinin resistance in malaria parasites. Science 367, 51–59, doi:10.1126/science.aax4735 (2020).

29 Mok, S. et al. Drug resistance. Population transcriptomics of human malaria parasites reveals the mechanism of artemisinin resistance. Science 347, 431–435, doi:10.1126/science.1260403 (2015).

30 Tilley, L., Straimer, J., Gnadig, N. F., Ralph, S. A. & Fidock, D. A. Artemisinin action and resistance in Plasmodium falciparum. Trends Parasitol 32, 682–696, doi:10.1016/j.pt.2016.05.010 (2016).

31 Gmp, W. Updated WHO policy recommendation: single dose primaquine as a gametocytocide in Plasmodium falciparum malaria. Geneva: WHO (2012).

32 Cottrell, G. et al. Emergence of resistance to atovaquone-proguanil in malaria parasites: insights from computational modeling and clinical case reports. Antimicrob Agents Chemother 58, 4504–4514, doi:10.1128/AAC.02550-13 (2014).

33 Staines, H. M. et al. Clinical implications of Plasmodium resistance to atovaquone. Antimicrob Agents Chemother 73, 581–595, doi:10.1093/jac/dkx431 (2018).

34 Goodman, C. D. et al. Parasites resistant to the antimalarial atovaquone fail to transmit by mosquitoes. Science 352, 349–353, doi:10.1126/science.aad9279 (2016).

35 Hogh, B. et al. The differing impact of chloroquine and pyrimethamine/sulfadoxine upon the infectivity of malaria species to the mosquito vector. Am J Trop Med Hyg 58, 176–182, doi:10.4269/ajtmh.1998.58.176 (1998).

36 Ecker, A., Lakshmanan, V., Sinnis, P., Coppens, I. & Fidock, D. A. Evidence that mutant PfCRT facilitates the transmission to mosquitoes of chloroquine-treated Plasmodium gametocytes. J Infect Dis 203, 228–236, doi:10.1093/infdis/jiq036 (2011).

37 Mharakurwa, S. et al. Malaria antifolate resistance with contrasting Plasmodium falciparum dihydrofolate reductase (DHFR) polymorphisms in humans and Anopheles mosquitoes. Proc Natl Acad Sci U S A 108, 18796–18801, doi:10.1073/pnas.1116162108 (2011).

38 Lambrechts, L., Halbert, J., Durand, P., Gouagna, L. C. & Koella, J. C. Host genotype by parasite genotype interactions underlying the resistance of anopheline mosquitoes to Plasmodium falciparum. Malar J 4, 3, doi:10.1186/1475-2875-4-3 (2005).

39 Hallett, R. L. et al. Chloroquine/sulphadoxine-pyrimethamine for gambian children with malaria: transmission to mosquitoes of multidrug-resistant Plasmodium falciparum. PLoS Clin Trials 1, e15, doi:10.1371/journal.pctr.0010015 (2006).

40 Targett, G. et al. Artesunate reduces but does not prevent posttreatment transmission of Plasmodium falciparum to Anopheles gambiae. J Infect Dis 183, 1254–1259, doi:10.1086/319689 (2001).

41 Gouagna, L. C. et al. Genetic variation in human HBB is associated with Plasmodium falciparum transmission. Nat Genet 42, 328–331, doi:10.1038/ng.554 (2010).

42 Joice, R. et al. Inferring developmental stage composition from gene expression in human malaria. PLoS Comput Biol 9, e1003392, doi:10.1371/journal.pcbi.1003392 (2013).

43 Graves, P. M., Carter, R., Burkot, T. R., Quakyi, I. A. & Kumar, N. Antibodies to Plasmodium falciparum gamete surface antigens in Papua New Guinea sera. Parasite Immunol 10, 209–218, doi:10.1111/j.1365-3024.1988.tb00215.x (1988).

44 Graves, P. M., Doubrovsky, A., Sattabongkot, J. & Battistutta, D. Human antibody responses to epitopes on the Plasmodium falciparum gametocyte antigen PFS 48/45 and their relationship to infectivity of gametocyte carriers. Am J Trop Med Hyg 46, 711–719, doi:10.4269/ajtmh.1992.46.711 (1992).

45 Usui, M. et al. Plasmodium falciparum sexual differentiation in malaria patients is associated with host factors and GDV1-dependent genes. Nature Commun 10, 2140, doi:10.1038/s41467-019-10172-6 (2019).

46 Bousema, T. et al. Human immune responses that reduce the transmission of Plasmodium falciparum in African populations. Int J Parasitol 41, 293–300, doi:10.1016/j.ijpara.2010.09.008 (2011).

47 Young, J. A. et al. The Plasmodium falciparum sexual development transcriptome: a microarray analysis using ontology-based pattern identification. Mol Biochem Parasitol 143, 67–79, doi:10.1016/j.molbiopara.2005.05.007 (2005).

48 Rockett, R. J. et al. A real-time, quantitative PCR method using hydrolysis probes for the monitoring of Plasmodium falciparum load in experimentally infected human volunteers. Malar J 10, 48, doi:10.1186/1475-2875-10-48 (2011).

49 Wang, C. Y. T. et al. Assays for quantification of male and female gametocytes in human blood by qRT-PCR in the absence of pure sex-specific gametocyte standards. Malar J 19, 218, doi:10.1186/s12936-020-03291-9 (2020).

50 Tadesse, F. G. et al. Molecular markers for sensitive detection of Plasmodium falciparum asexual stage parasites and their application in a malaria clinical trial. Am J Trop Med Hyg 97, 188–198, doi:10.4269/ajtmh.16-0893 (2017).

51 Bancells, C. et al. Revisiting the initial steps of sexual development in the malaria parasite Plasmodium falciparum. Nat Microbiol 4, 144–154, doi:10.1038/s41564-018-0291-7 (2019).

52 Farid, R., Dixon, M. W., Tilley, L. & McCarthy, J. S. Initiation of gametocytogenesis at very low parasite density in Plasmodium falciparum infection. J Infect Dis 215, 1167–1174, doi:10.1093/infdis/jix035 (2017).

53 Wang, C. Y. T. et al. Assessing Plasmodium falciparum transmission in mosquito-feeding assays using quantitative PCR. Malar J 17, 249, doi:10.1186/s12936-018-2382-6 (2018).

