## Supplemental material for "*Plasmodium falciparum* artemisinin-resistant K13 mutations confer a sexual-stage transmission advantage that can be overcome with atovaquone-proguanil"

**Supplementary Material**

**Supplementary Tables** ........ **3**

Supplementary Table 1. List of markers for male and female gene expression used in analyses of isolates collected in the TRAC I study........ 6

Supplementary Table 2. Gametocyte transcript levels per isolate collected in the TRAC I study (xls). ........ 3

Supplementary Table 3. Day of treatment in the K13 vs 3D7 clinical trial. ........ 4

Supplementary Table 4. Regression analysis between pre-treatment total parasitaemia and total gametocytemia for ART-resistant (ARTr) and ART-sensitive (ARTs) infected participants. ........ 3

Supplementary Table 5. Male to female gametocyte ratio in ART-resistant (ARTr) and ART-sensitive infected volunteers (ARTs). ........ 5

**Supplementary Figures** ........ **7**

Supplementary Figure 2. Correlation between total parasitaemia area under the curve (AUC) and total gametocytemia ($log_{10}$-transformed) in the K13 vs 3D7 clinical trial. ... 8

### Supplementary Tables

**Supplementary Table 1. Gametocyte transcript levels per isolate collected in the TRAC I study (xls).**

**Supplementary Table 2. Regression analysis between pre-treatment total parasitaemia and total gametocytemia for ART-resistant (ARTr) and ART-sensitive (ARTs) infected participants.**

| Strain | Relationship | Slope (95% CI) | *p* (model) | Adjusted $R^2$ |
|---|---|---|---|---|
| ARTr | Log total parasitaemia pre-artesunate AUC versus log-transformed total gametocytemia AUC, 12 days post-artesunate | 2.23 (0.74-1.53) | 0.005 | 0.52 |
| ARTs | Log total parasitaemia pre-artesunate AUC versus log-transformed total gametocytemia AUC, 12 days post-artesunate | 1.34 (0.27-2.04) | 0.510 | 0.06 |

ARTr: ART-resistant strain, Cam3.II$^{R539T}$ and ARTs: ART-sensitive strain, 3D7

**Supplementary Table 3. Day of treatment in the K13 vs 3D7 clinical trial.**

| K13 vs 3D7 clinical trial | *P. falciparum strain* | Volunteer | Artesunate (2 mg/kg) | Piperaquine (960 mg/kg) | Atovaquone-Proguanil (250/100 mg/kg) | Primaquine (45 mg/kg) |
|---|---|---|---|---|---|---|
| Cohort 1 | Cam3.II$^{R539T}$ | ARTr_1 | D9 | D11 | D20 | D28 |
| | Cam3.II$^{R539T}$ | ARTr_2 | D9 | D11 | D20 | D28 |
| | Cam3.II$^{R539T}$ | ARTr_3 | D9 | D11 | D16 | D28 |
| | 3D7 | ARTs_1 | D9 | D17 | D27 | D35 |
| | 3D7 | ARTs_2 | D9 | D20 | D27 | D31 |
| | 3D7 | ARTs_3 | D9 | D17 | D27 | D35 |
| Cohort 2 | Cam3.II$^{R539T}$ | ARTr_4 | D9 | D11 | D28 | D23 |
| | Cam3.II$^{R539T}$ | ARTr_5 | D9 | D11 | D21 | D23 |
| | Cam3.II$^{R539T}$ | ARTr_6 | D9 | D11 | D19 | D23 |
| | Cam3.II$^{R539T}$ | ARTr_7 | D9 | D11 | D28 | D23 |
| | Cam3.II$^{R539T}$ | ARTr_8 | D9 | D11 | D28 | D23 |
| | Cam3.II$^{R539T}$ | ARTr_9 | D9 | D11 | D28 | D23 |
| | Cam3.II$^{R539T}$ | ARTr_10 | D9 | D11 | D28 | D23 |
| | 3D7 | ARTs_4 | D9 | D19 | D28 | D23 |
| | 3D7 | ARTs_5 | D9 | D17 | D28 | D23 |
| | 3D7 | ARTs_6 | D9 | D17 | D28 | D23 |
| Cohort 3 | Cam3.II$^{R539T}$ | ARTr_11 | D9 | D11 | D21 | D24 |
| | Cam3.II$^{R539T}$ | ARTr_12 | D9 | D11 | D21 | D24 |
| | Cam3.II$^{R539T}$ | ARTr_13 | D9 | D11 | D21 | D24 |
| | 3D7 | ARTs_7 | D9 | D15 | D21 | D24 |
| | 3D7 | ARTs_8 | D9 | D15 | D21 | D24 |
| | 3D7 | ARTs_9 | D9 | D15 | D21 | D24 |

Study day (D); ART-resistant parasites (ARTr); ART-sensitive parasites (ARTs).

**Supplementary Table 4. Male to female gametocyte ratio in ART-resistant (ARTr) and ART-sensitive (ARTs) infected volunteers.**

Two sample *t*-test performed on the log transformed male to female gametocyte ratio and back-transformed to present as mean (95% Confidence interval).

| **Period** | **ART-resistant** | | **ART-sensitive** | | **Two Sample *t*-test** |
|---|---|---|---|---|---|
| | **n** | **Mean (95% CI)** | **n** | **Mean (95% CI)** | ***p*** |
| Overall | 68 | 0.200 (0.159 - 0.252) | 18 | 0.096 (0.062-0.148) | 0.004 |
| <20 days | 33 | 0.146 (0.111 - 0.194) | 5 | 0.090 (0.022 - 0.368) | 0.231 |
| 20-22 days | 16 | 0.220 (0.142 - 0.339) | 6 | 0.087 (0.034 - 0.224) | 0.031 |
| >22 days | 19 | 0.319 (0.187 - 0.546) | 7 | 0.109 (0.053 - 0.223) | 0.028 |

**Supplementary Table 5. List of markers for male and female gene expression used in analyses of isolates collected in the TRAC I study.**

| *PlasmoDB Gene ID* | Gene description | Gametocyte-specific |
|---|---|---|
| ***PF3D7_1031000*** | **P25, 25-kDa ookinete surface antigen precursor (Pfs25)** | **Female** |
| *PF3D7_1246200* | Actin I | Female |
| *PF3D7_0816800* | DMC1, meiotic recombination protein, putative | Female |
| *PF3D7_1143100* | AP2-O, AP2 domain transcription factor, putative | Female |
| *PF3D7_0621400* | Pf77 | Female |
| *PF3D7_1128300* | PFK11, ATP-dependent 6-phosphofructokinase | Female |
| *PF3D7_1475500* | CCp1, LCCL domain-containing protein | Female |
| *PF3D7_1407000* | CCp3, LCCL domain-containing protein | Female |
| *PF3D7_1426500* | ABCG2, ABC transporter G family member 2 | Female |
| *PF3D7_1250100* | G377, osmiophilic body protein G377 | Female |
| *PF3D7_1346800* | P47, 6-cysteine protein | Female |
| *PF3D7_1107800* | ApiAP2, AP2 domain transcription factor, putative | Female |
| *PF3D7_0719200* | NEK4, NIMA-related kinase 4 | Female |
| *PF3D7_1455800* | CCp2, LCCL domain-containing protein | Female |
| *PF3D7_1113900* | MAPK2, mitogen-activated protein kinase 2 | Male |
| *PF3D7_1311100* | Meiosis-specific nuclear structural protein 1, putative | Male |
| ***PF3D7_1325200*** | **Lactate dehydrogenase, putative (MGET)** | **Male** |
| ***PF**3D7_1122900* | Dynein heavy chain, putative | Male |
| *PF3D7_1114000* | Dynein light chain Tctex-type, putative | Male |
| *PF3D7_1014200* | HAP2, gamete fusion factor, putative | Male |

The two genes also used in qRT-PCR in the K13 vs 3D7 study are highlighted in bold.

### Supplementary Figures

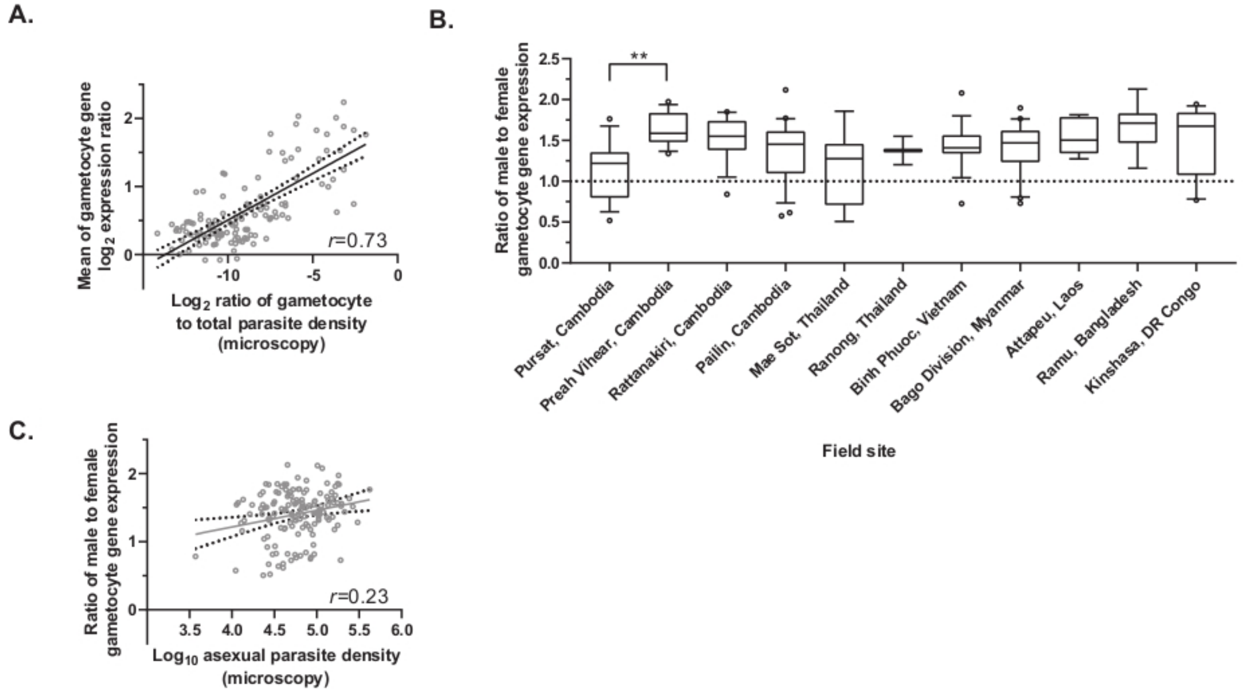

**Supplementary Figure 1. Gametocyte dynamics and parasite density correlation in symptomatic clinical samples collected from the TRAC I study in 2011-2013. A)** Scatter plot showing a positive correlation between gametocyte gene expression levels and ratios of gametocyte to total parasite density derived from microscopy counts (n=126; taking samples with gametocytes observed from microscopy). The dotted lines represent the 95% confidence interval bands of the linear regression model. **B)** Box and whisker plots of the median and 10-90 percentile show variation in male to female gametocyte ratios in all field sites (n=139; taking samples where the average male or female gametocyte transcript levels are higher than the asexual reference pool by ≥1.42-fold). ** adjusted p<0.01. **C)** Scatter plot showing positive association between ratio of male to female gametocytes and asexual parasite density (n=139; taking samples where the average male or female gametocyte transcript levels were higher than the asexual reference pool by ≥1.42-fold). The dotted lines represent the 95% confidence interval bands of the linear regression model.

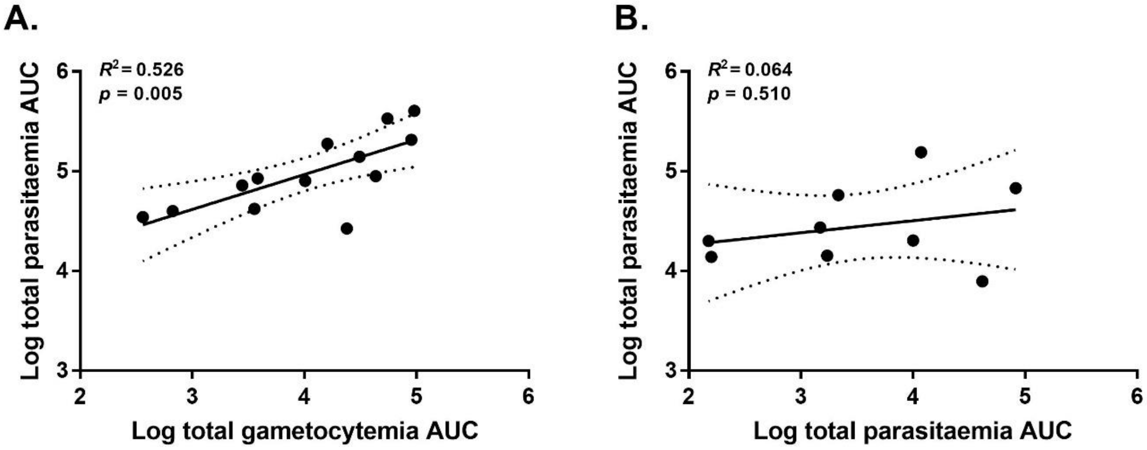

**Supplementary Figure 2. Correlation between total parasitaemia area under the curve (AUC) and total gametocytemia ($log_{10}$-transformed) in the K13 vs. 3D7 clinical trial. A)** A linear regression showed a moderate correlation between asexual AUC and total gametocytemia AUC in ART-resistant infected volunteers **B)** No correlation between asexual AUC and total gametocytemia AUC was found in ART-sensitive infected volunteers. 95% confidence interval (dotted line) and mean (solid line).

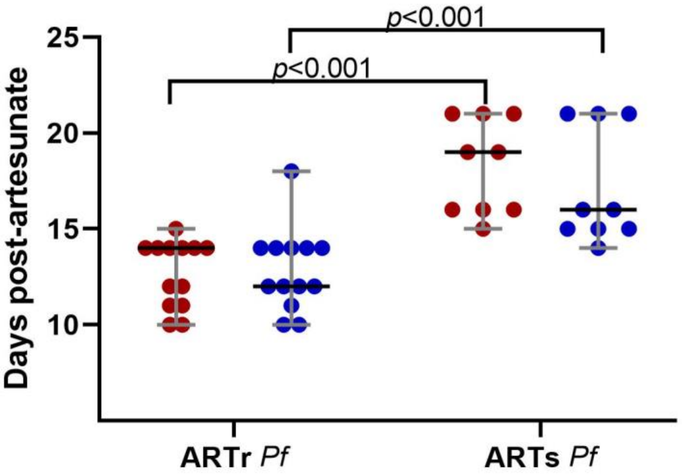

**Supplementary Figure 3. Day post-artesunate when peak gametocytemia was reached.** Peak gametocytaemia on ART-resistant infected volunteers (ARTr) was reached on a median of 12-14 days post-artesunate (range: 10-18 days), and in ART-sensitive infected volunteers (ARTs) 16-19 days post artesunate (range: 14-21 days); Mann-Whitney *U* test; $p > 0.001$. Female and male gametocytes were quantified by qRT-PCR targeting *pfs25* and *pfMGET* respectively.

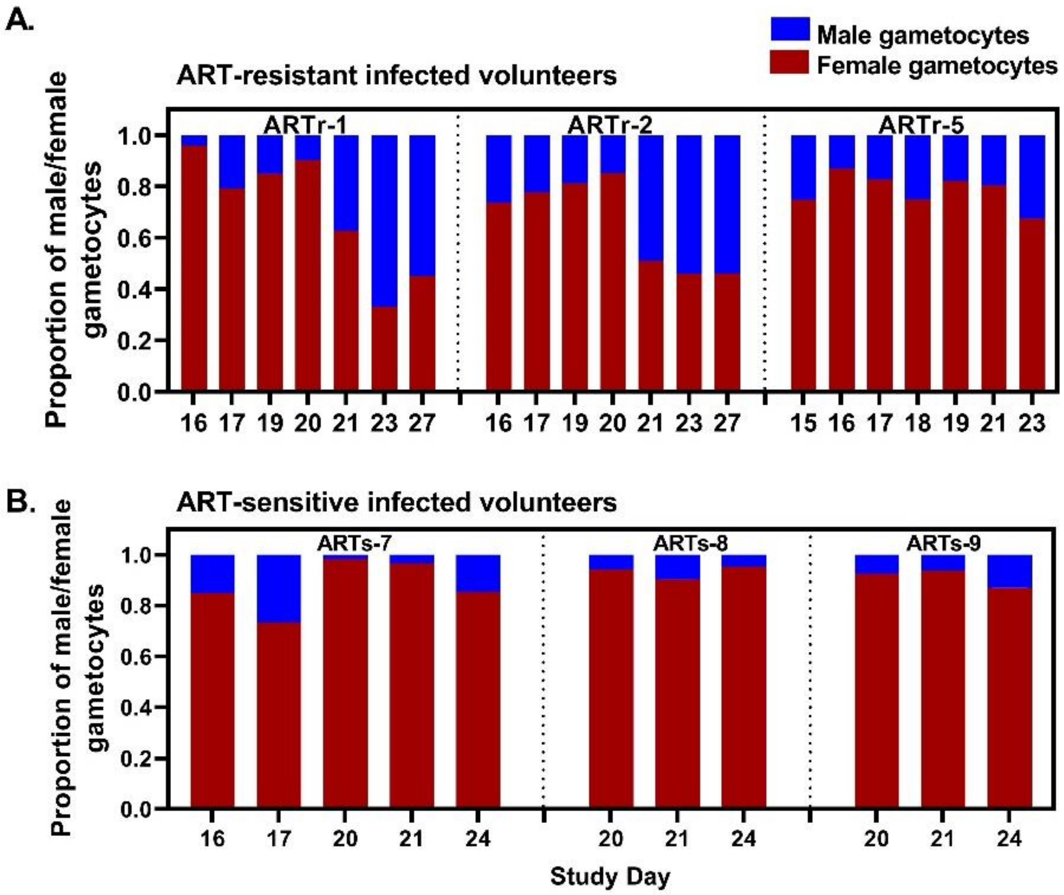

**Figure 4. Male and female gametocyte proportions varied over time in the K13 vs 3D7 clinical trial. A)** Proportion of male or female gametocytes over time for three ART-resistant infected volunteers before treatment with primaquine. **B)** Proportion of male or female gametocytes over time for three ART-sensitive infected volunteers before treatment with primaquine. Gametocyte development was monitored by qRT-PCR for *pfs25* (female gametocytes) and *pfMGET* (male gametocytes) over time.

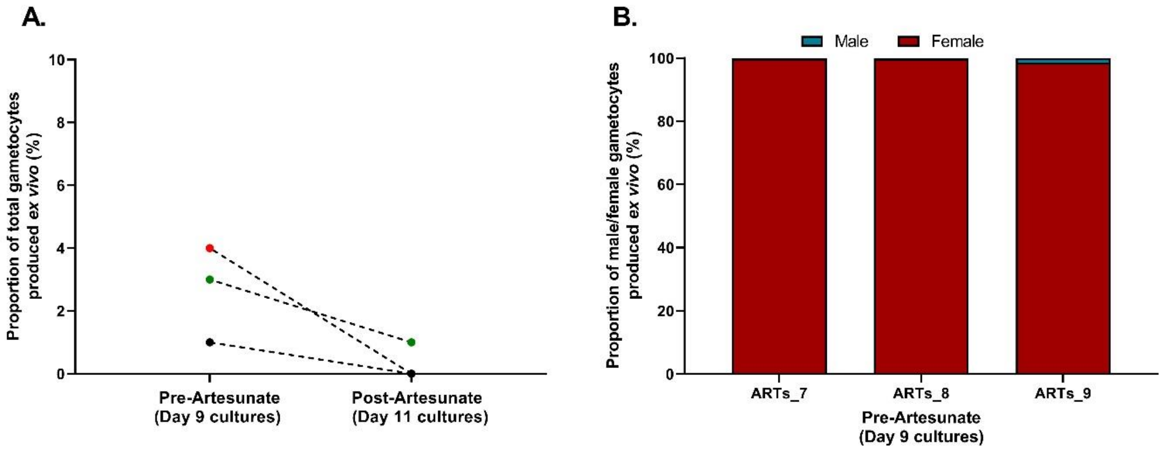

**Supplementary Figure 5. *Ex vivo* ART-sensitive cultures from samples taken post-artesunate treatment did not develop gametocytes. A)** Proportion of total gametocytes (male and female) per total asexual parasitaemia developed after 15 days of *ex vivo* culture from samples taken pre and post artesunate. **B)** Bar-plots showing the sex composition of the *ex vivo* cultures 15 days after sample collection on Day 9. Female and male gametocytes were quantified by qRT-PCR targeting *pfs25* and *pfMGET* respectively, total parasitaemia was quantified by qPCR targeting 18S-rRNA.

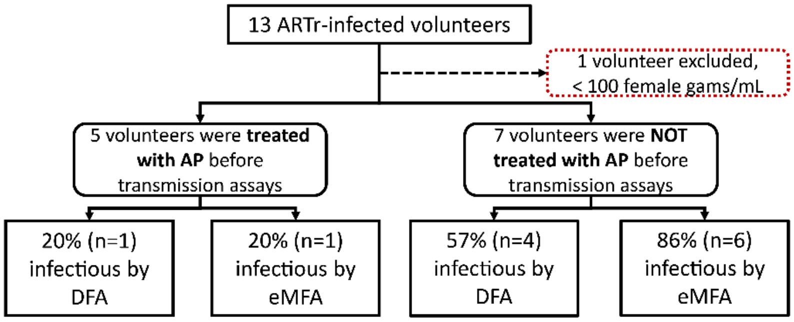

**Supplementary Figure 6. ART-resistant infected volunteers (ARTr) enrolled in transmission studies.** One of the 13 ART-resistant infected volunteers was excluded from the analysis because of low gametocytemia before transmission assays (less than 100 gametocytes/mL). Five volunteers were rescued with atovaquone-proguanil (AP) before transmission assays, and seven did not require AP treatment before transmission assays.

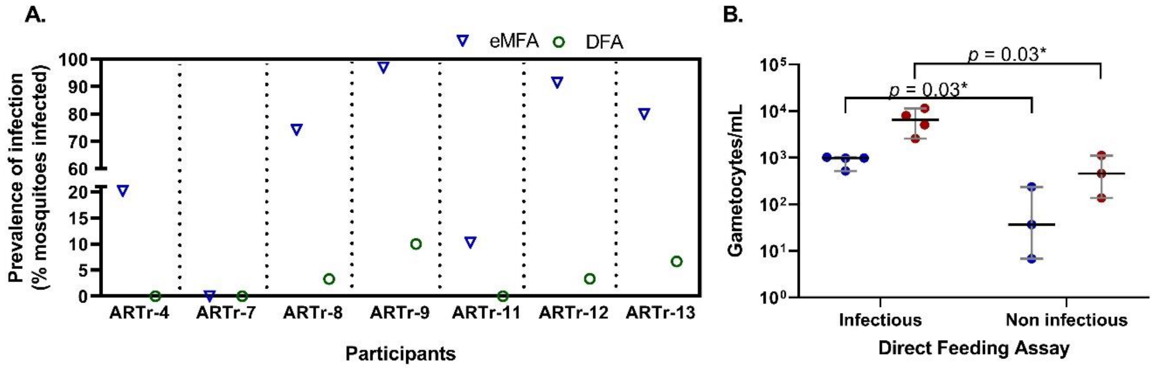

**Supplementary Figure 7. Transmissibility of ART-resistant infected volunteers (ARTr). A)** Proportion of infected midguts by enriched Membrane Feeding Assay (eMFA, blue triangles) and by Direct Feeding Assay (DFA, green circles); ARTr-4 –9 assessed on Day 23, ARTr-11 -13 on Day 21. **B)** Gametocytemia at time of DFA in infectious vs. non-infectious volunteers by DFA. Median female and male gametocytemia was significantly higher at the time of the assay in infectious volunteers versus non-infections by DFA. Infectious volunteers: 456 female gams/mL (range: 137-1,115) and 37 male gams/mL (range:7-237) vs. non-infectious volunteers: 6,543 female gams/mL (range: 2,569-11,450) and 983 male gams/mL (range: 518-1029); Mann-Whitney *U* test; *p*=0.03. Female and male gametocytes were quantified by qRT-PCR targeting *pfs25* and *pfMGET* respectively.

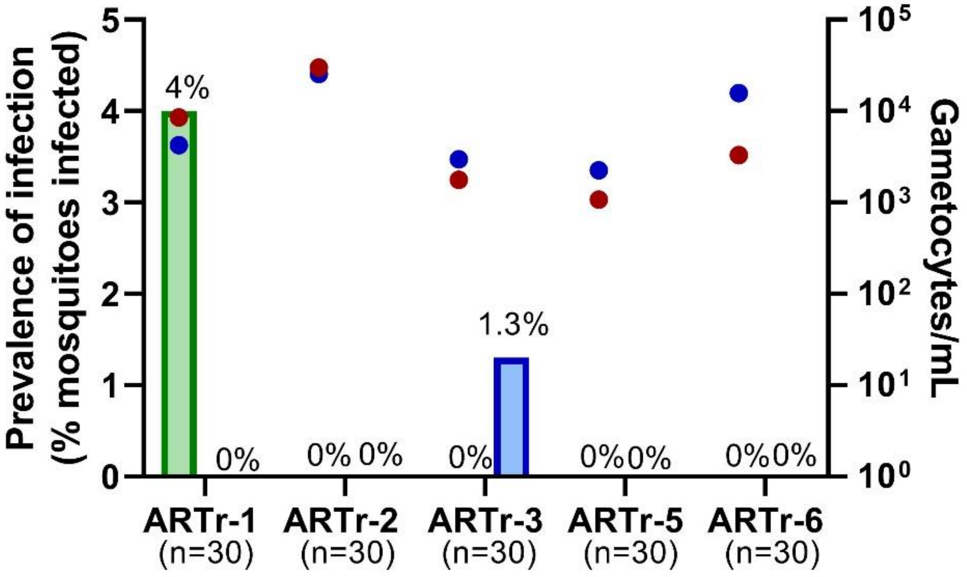

**Supplementary Figure 8. Transmissibility of ART-resistant (ARTr) infected volunteers rescued with atovaquone-proguanil (AP) before transmission assays on Day 23.** Infection rates were very low or not present. Proportion of infected midguts by Direct Feeding Assay (DFA, green bars) and by enriched Membrane Feeding Assay (eMFA, blue bars). Red circles indicate female gametocytemia (gametocytes/mL) and blue circles indicate male gametocytemia (gametocytes/mL). Female and male gametocytes were quantified by qRT-PCR targeting *pfs25* and *pfMGET* respectively, at the time of DFA.
